# Effects of a DPP-4 inhibitor and RAS blockade on clinical outcomes of patients with diabetes and COVID-19

**DOI:** 10.1101/2020.05.20.20108555

**Authors:** Sang Youl Rhee, Jeongwoo Lee, Hyewon Nam, Dae-Sung Kyoung, Dae Jung Kim

**Affiliations:** Department of Endocrinology and Metabolism, Kyung Hee University School of Medicine, Seoul, Korea; Data Science team, Hanmi Pharm. Co., Ltd., Seoul, Korea; Department of Endocrinology and Metabolism, Ajou University School of Medicine, Suwon, Korea

**Keywords:** Diabetes Mellitus, Dipeptidyl Peptidase-4, Angiotensin-Converting Enzyme 2, COVID-19, COVID-19 drug treatment

## Abstract

**Background:** Dipeptidyl peptidase-4 inhibitor (DPP-4i) and renin–angiotensin system (RAS) blockade are reported to affect the clinical course of coronavirus disease 2019 (COVID-19) in patients with diabetes mellitus (DM). However, the effectiveness of these drugs in large populations is unclear.

**Subjects and Methods:** As of May 2020, data analysis was conducted on all subjects who could confirm their history of claims related to COVID-19 in the National Health Review and Assessment Service database in Korea. Using the COVID-19 and claims data of the past 5 years, we compared the short-term prognosis of COVID-19 infection according to the use of DPP-4i and RAS blockade.

**Results:** Totally, data of 67850 subjects were accessible. Of these, 5080 were confirmed COVID-19. Among these, 832 subjects with DM were selected for analysis in this study. Among the subjects, 263 (31.6%) and 327 (39.3%) were DPP-4i and RAS blockade users, respectively. Thirty-four subjects (4.09%) received intensive care or died. The adjusted odds ratio for severe treatment among DPP-4i users was 0.362 [95% confidence interval (CI), 0.135–0.971], and that for RAS blockade users was 0.599 (95% CI, 0.251–1.431). No synergy was observed for subjects using both drugs.

**Conclusion:** This population-based study suggests that DPP-4i is significantly associated with a better clinical outcome of patients with COVID-19. However, the effect of RAS blockade is not significant.

## Introduction

People with risk factors for cardiovascular diseases such as diabetes mellitus (DM) and hypertension have a higher risk of COVID-19 infection than the general population and generally exhibit poor prognosis^1-5^. There is an urgent need to develop evidence-based treatment methods to effectively prevent and manage COVID-19 in people with chronic diseases. However, it is not easy to provide reliable evidence within a short period in a worldwide pandemic situation.

Recently, it has been suggested that dipeptidyl peptidase-4 (DPP-4) and angiotensin-converting enzyme 2 (ACE2) may be parts of the receptor proteins of the novel coronavirus and may have a significant impact on the clinical course of COVID-19^6,7^. Their molecular pathways are well-known to play important roles in glucose homeostasis, cardiovascular hemodynamics, and physiologic responses associated with inflammatory cascades^8-12^. Therefore, DPP-4 inhibitor (DPP-4i) and renin-angiotensin system (RAS) blockade, which are widely used in general practice, are likely to have a significant impact on the clinical course of COVID-19 in patients with chronic diseases^13,14^.

The evidence that DPP-4i and RAS blockade significantly affect the clinical course of COVID-19 infection is unclear. Therefore, it is generally recommended that COVID-19 patients with chronic diseases either not use or discontinue these drugs^15^. However, the clinical significance of DPP-4i and RAS blockade for COVID-19 in the absence of effective treatment or preventive measures other than symptomatic treatment can be very important.

Hence, we utilized the entire claims data related to COVID-19 from Korea’s national medical insurance database, and investigated the effects of DPP-4i and RAS blockade, which are commonly administered to patients with DM, on the short-term clinical outcomes of COVID-19.

## Subjects and Methods

Health Insurance Review & Assessment Service (HIRA) claims database

The Korean national health insurance is a single healthcare insurance system that covers approximately 97% of the total Korean population; the remaining 3% are “medical protection” beneficiaries. Information on individuals’ utilization of medical facilities, prescription records, and diagnostic codes configured in the format of International Classification of Diseases, 10th revision (ICD-10) is recorded in the HIRA database^16^. This database is considered representative of the Korean population and is used in research through anonymization and de-identification.

### Study subjects

Currently, in Korea, medication usage data for the past 5 years for those who have claimed a confirmatory test of COVID-19 are managed as a separate database for research. To protect personal information, the export of raw data is strictly prohibited by law, and research is being performed in a way that provides de-identified results when researchers submit program codes for analysis^17^. As of May 17, 2020, the total number of Korean COVID-19 claims released to researchers was 67850 (Figure 1). Of these, 5080 and 832 were confirmed cases of COVID-19 and COVID-19 with DM, respectively.

**Figure 1.**
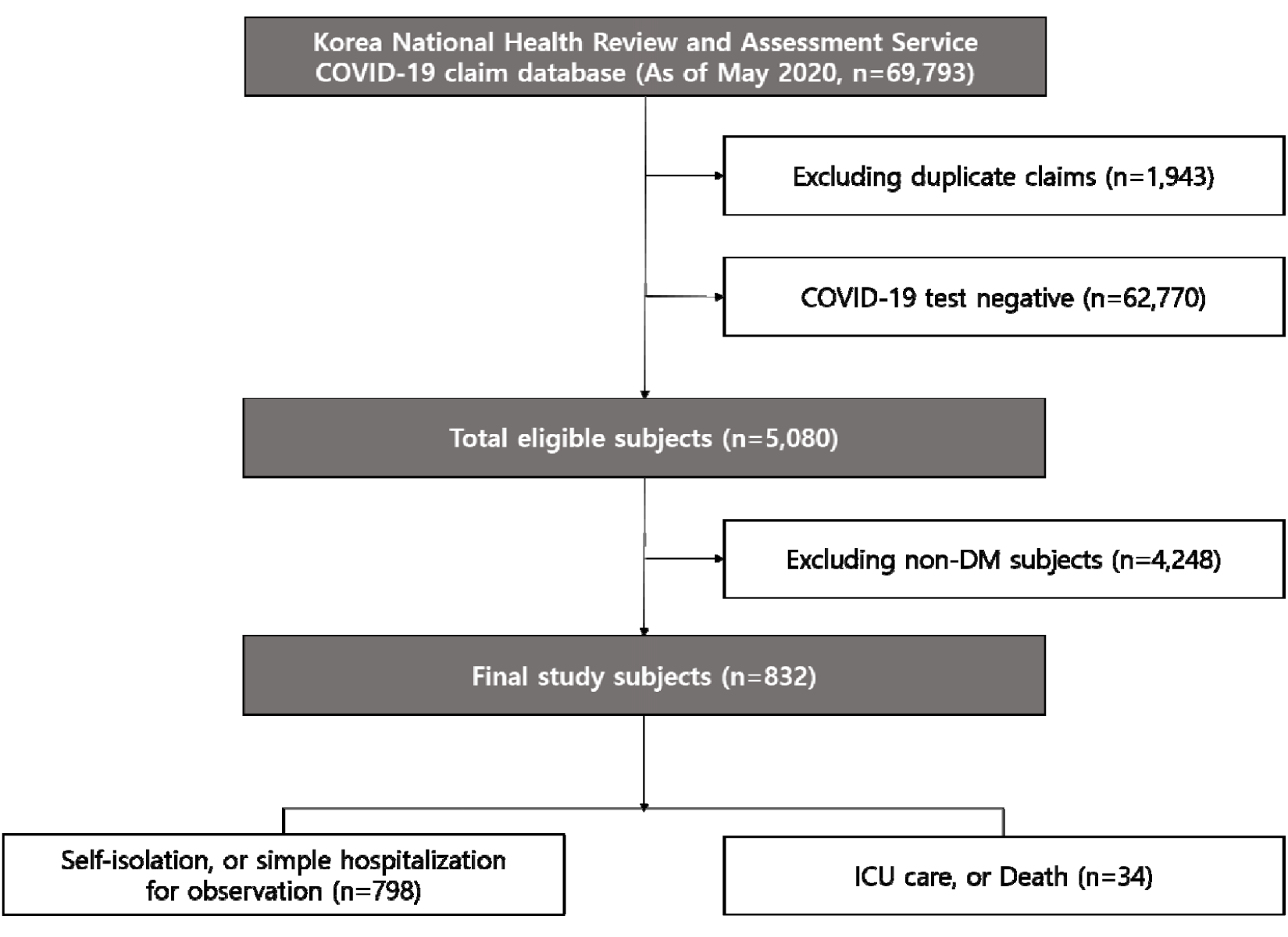
Flow chart of the selection of study subjects.

### Definition of COVID-19 infection, clinical status, and other clinical variables

The clinical variables that can be identified in Korea’s COVID-19 claim database are the subject’s age, sex, diagnostic code, medication prescription, outpatient care, hospitalization, critical care, and death. COVID-19 diagnosis was defined as ICD-10 diagnosis codes B34.2, B97.2, U18, U18.1, or U07.x. Intensive care was defined as a procedure code for endotracheal intubation or mechanical ventilation, or the charge of an intensive care unit management fee. We analyzed subjects according to a mild case of self-isolation or simple hospitalization, or a severe/lethal case of death or intensive care.

The subject’s comorbidity was defined as a diagnostic code. DM was defined as E10.x, E11.x, E12.x, E13.x, or E14.x, hypertension as |10.x, |11.x, |12.x, |13.x, or |15.x, dyslipidemia as E78.x, cardiovascular disease as |20.x, |21.x, |22.x, |23.x, |24.x, or |25.x, and cerebrovascular disease as |61.x, |62.x, |63.x, or |64.x. Chronic kidney disease was defined as N18.x, asthma as J45.x or J46.x, and chronic obstructive pulmonary disease (COPD) as J44.x. The subject’s medication use was assessed based on the initial date of COVID-19 diagnosis. If the prescription was confirmed within 180 days of diagnosis and was for at least 90 days, the subject was defined as a user of a specific drug. Thereafter, differences in characteristics and clinical status were compared according to whether the subject used DPP-4i and/or RAS blockade. We also investigated the possibility of synergy during the combined use of these two drugs.

### Statistical analysis

Basic characteristics of subjects were expressed as mean ± standard deviation for continuous variables in each subgroup and as number and percentage for categorical variables. The Chi-square test, Fisher’s exact test, independent Student’s t-test, and one-way analysis of variance were used to compare the clinical characteristics of the subjects. Logistic regression analysis was used to adjust various clinical variables. Statistical analyses were performed using SAS version 9.4 (SAS Institute, Cary, NC, US), and *p<* 0.05 was considered statistically significant.

### Ethics statement

This study was approved by the Institutional Review Board of Kyung Hee University Hospital (no. KHUH 2020-04-067). The requirement for informed consent was waived by the institutional review board because de-identified information was used for the analyses.

## Results

### Clinical characteristics with or without use of DPP-4i

Clinical characteristics of subjects were compared according to whether DPP-4i was used (Table 1). The mean age of DPP-4i users was 63.69 ± 12.19 years, which was significantly higher than that of non-users. Of these, males were 56.65%, which was not significantly different from the percentage among non-users. Hypertension and dyslipidemia were shown by 74.90% and 93.92% of the subjects, respectively. There were no significant differences in the prevalence of other comorbidities. In terms of medication, DPP-4i users had significantly higher rates of metformin, sulfonylurea, thiazolidinedione, RAS blockade, diuretics, statin, and fibrate usage. However, the usage rate of sodium–glucose cotransporter-2 inhibitor (SGLT2i) was significantly higher in non-users.

**Table 1.** Clinical characteristics according to the use of DPP-4 inhibitors in Korean COVID-19 patients with diabetes.

|  | No DPP-4 inhibitor<br>user (n=569) | DPP-4 inhibitor user<br>(n=263) | *p* |
| --- | --- | --- | --- |
| Age (years) | 60.99±16.90 | 63.69±12.19 | <0.0001 |
| <40 (n, %) | 65 (11.42) | 4 (1.52) |  |
| 40 to <50 (n, %) | 74 (13.01) | 29 (11.03) |  |
| 50 to <60 (n, %) | 114 (20.04) | 61 (23.19) | <0.0001 |
| 60 to <70 (n, %) | 125 (21.97) | 90 (34.22) |  |
| 70 to <80 (n, %) | 112 (19.68) | 55 (20.91) |  |
| ≥80 (n, %) | 79 (13.88) | 24 (9.13) |  |
| Male (n, %) | 296 (52.02) | 149 (56.65) | 0.2129 |
| Hypertension (n, %) | 371 (65.20) | 197 (74.90) | 0.0052 |
| Dyslipidemia (n, %) | 483 (84.89) | 247 (93.92) | 0.0002 |
| Cardiovascular disease (n, %) | 159 (27.94) | 68 (25.86) | 0.5295 |
| Cerebrovascular disease (n, %) | 64 (11.25) | 41 (15.59) | 0.0795 |
| Chronic kidney disease (n, %) | 104 (18.28) | 58 (22.05) | 0.2010 |
| Asthma (n, %) | 166 (29.17) | 73 (27.76) | 0.6744 |
| COPD (n, %) | 51 (8.96) | 21 (7.98) | 0.6407 |
| Cancer (n, %) | 136 (23.90) | 48 (18.25) | 0.0679 |
| Medication |  |  |  |
| Metformin (n, %) | 86 (15.11) | 183 (69.58) | <0.0001 |
| Sulfonylurea (n, %) | 30 (5.27) | 100 (38.02) | <0.0001 |
| Meglitinide (n, %) | 5 (0.88) | 2 (0.76) | 0.8621 |
| TZD (n, %) | 8 (1.41) | 33 (12.55) | <0.0001 |
| DPP-4 inhibitor (n, %) | 0 (0.00) | 263 (100.00) | NA |
| SGLT2 inhibitor (n, %) | 18 (3.16) | 1 (0.38) | 0.0125 |
| AGI (n, %) | 6 (1.05) | 1 (0.38) | 0.3222 |
| Insulin (n, %) | 1 (0.18) | 1 (0.38) | 0.5755 |
| RAS blockade (n, %) | 197 (34.62) | 130 (49.43) | <0.0001 |
| ACEi (n, %) | 18 (3.16) | 6 (2.28) | 0.4797 |
| ARB (n, %) | 182 (31.99) | 124 (47.15) | <0.0001 |
| Beta blocker (n, %) | 92 (16.17) | 46 (17.49) | 0.6337 |
| Diuretics (n, %) | 65 (11.42) | 46 (17.49) | 0.0167 |
| CCB (n, %) | 171 (30.05) | 88 (33.46) | 0.3237 |
| Statin (n, %) | 207 (36.38) | 159 (60.46) | <0.0001 |
| Fibrate (n, %) | 15 (2.64) | 17 (6.46) | 0.0076 |

COPD indicates chronic obstructive pulmonary disease; TZD, thiazolidinedione; DPP-4, dipeptidyl peptidase-4; SGLT2, sodium glucose cotransporter 2; AGI, alpha glucosidase inhibitor; RAS, renin-angiotensin system; ACEi, angiotensin converting enzyme inhibitor; ARB, angiotensin receptor blocker; CCB, calcium channel blocker, NA, not applicable.

### Clinical characteristics with or without the use of RAS blockade

Clinical characteristics of subjects were compared according to whether RAS blockade was used (Table 2). RAS-blockade users had a mean age of 64.85 ± 13.23 years, which was significantly higher than that of non-users, and 56.57% of them were male. Most RAS-blockade users had dyslipidemia, and the prevalence of cardiovascular and chronic kidney diseases among them was significantly higher than that among non-users. With regard to medication, the usage frequency of metformin, sulfonylurea, thiazolidinedione, DPP-4i, diuretic, calcium-channel blocker, and statin was significantly higher among RAS-blockade users than among nonusers.

**Table 2.** Clinical characteristics according to the use of RAS blockade in Korean COVID-19 patients with diabetes.

|  | No RAS blockade user<br>(n=505) | RAS blockade user<br>(n=327) | *p* |
| --- | --- | --- | --- |
| Age (years) | 59.90±16.70 | 64.85±13.23 | <0.0001 |
| <40 (n, %) | 60 (11.88) | 9 (2.75) | 0.0001 |
| 40 to <50 (n, %) | 66 (13.07) | 37 (11.31) |  |
| 50 to <60 (n, %) | 108 (21.39) | 67 (20.49) |  |
| 60 to <70 (n, %) | 122 (24.16) | 93 (28.44) |  |
| 70 to <80 (n, %) | 92 (18.22) | 75 (22.94) |  |
| ≥80 (n, %) | 57 (11.29) | 46 (14.07) |  |
| Male (n, %) | 260 (51.49) | 185 (56.57) | 0.1505 |
| Hypertension (n, %) | 242 (47.92) | 326 (99.69) | <0.0001 |
| Dyslipidemia (n, %) | 427 (84.55) | 303 (92.66) | 0.0005 |
| Cardiovascular disease (n, %) | 125 (24.75) | 102 (31.19) | 0.0417 |
| Cerebrovascular disease (n, %) | 62 (12.28) | 43 (13.15) | 0.7112 |
| Chronic kidney disease (n, %) | 68 (13.47) | 94 (28.75) | <0.0001 |
| Asthma (n, %) | 151(29.90) | 88 (26.91) | 0.3519 |
| COPD (n, %) | 47 (9.31) | 25 (7.65) | 0.4051 |
| Cancer (n, %) | 118 (23.37) | 66 (20.18) | 0.2799 |
| Medication |  |  |  |
| Metformin (n, %) | 142 (28.12) | 127 (38.84) | 0.0012 |
| Sulfonylurea (n, %) | 63 (12.48) | 67 (20.49) | 0.0019 |
| Meglitinide (n, %) | 4 (0.79) | 3 (0.92) | 0.8467 |
| TZD (n, %) | 17 (3.37) | 24 (7.34) | 0.0097 |
| DPP-4 inhibitor (n, %) | 133 (26.34) | 130 (39.76) | <0.0001 |
| SGLT2 inhibitor (n, %) | 9 (1.78) | 10 (3.06) | 0.2288 |
| AGI (n, %) | 4 (0.79) | 3 (0.92) | 0.8467 |
| Insulin (n, %) | 1 (0.20) | 1 (0.31) | 0.7565 |
| RAS blockade (n, %) | 0 (0.00) | 327 (100.00) | NA |
| ACEi (n, %) | 0 (0.00) | 24 (7.34) | NA |
| ARB (n, %) | 0 (0.00) | 306 (93.58) | NA |
| Beta blocker (n, %) | 62 (12.28) | 76 (23.24) | <0.0001 |
| Diuretics (n, %) | 32 (6.34) | 79 (24.16) | <0.0001 |
| CCB (n, %) | 83 (16.44) | 176 (53.82) | <0.0001 |
| Statin (n, %) | 174 (34.46) | 192 (58.72) | <0.0001 |
| Fibrate (n, %) | 16 (3.17) | 16 (4.89) | 0.2064 |

COPD indicates chronic obstructive pulmonary disease; TZD, thiazolidinedione; DPP-4, dipeptidyl peptidase-4; SGLT2, sodium glucose cotransporter 2; AGI, alpha glucosidase inhibitor; RAS, renin-angiotensin system; ACEi, angiotensin converting enzyme inhibitor; ARB, angiotensin receptor blocker; CCB, calcium channel blocker, NA, not applicable.

### Differences in severity of COVID-19 infection upon use of DPP-4i and RAS blockade

The fractions of those who received intensive care or died were 3.42% and 4.39% among DPP-4i users and non-users, respectively (Table 3). The unadjusted odds ratio (OR) of these severe/lethal cases among DPP-4i users was 0.771 (95% confidence interval (CI), 0.355–1.676). However, the adjusted OR (aOR), considering age, sex, comorbidity, and medication, was 0.362 (95% CI, 0.135–0.971).

**Table 3.** Differences in COVID-19 related clinical status on the use of DPP-4 inhibitors and/or RAS blockade.

|  | total<br>case<br>(n) | mild<br>case<br>(n) | Intensive<br>care or<br>death (n) | Model 1* |  | Model 2** |  | Model 3*** |  | Model 4**** |  |
| --- | --- | --- | --- | --- | --- | --- | --- | --- | --- | --- | --- |
|  |  |  |  | OR | 95% CI | OR | 95% CI | OR | 95% CI | OR | 95% CI |
| No DPP-4i user | 569 | 544 | 25 | 1.00 (ref.) |  | 1.00 (ref.) |  | 1.00 (ref.) |  | 1.00 (ref.) |  |
| DPP-4i user | 263 | 254 | 9 | 0.771 | 0.355-1.676 | 0.758 | 0.344-1.671 | 0.651 | 0.283-1.497 | 0.362 | 0.135-0.971 |
| No RAS blockade user | 505 | 484 | 21 | 1.00 (ref.) |  | 1.00 (ref.) |  | 1.00 (ref.) |  | 1.00 (ref.) |  |
| RAS blockade user | 327 | 314 | 13 | 0.954 | 0.471-1.933 | 0.780 | 0.379-1.602 | 0.605 | 0.268-1.365 | 0.599 | 0.251-1.431 |
| No RAS, No DPP-4i user | 372 | 354 | 18 | 1.00 (ref.) |  | 1.00 (ref.) |  | 1.00 (ref.) |  | 1.00 (ref.) |  |
| RAS only user | 197 | 190 | 7 | 0.725 | 0.297-1.766 | 0.591 | 0.239-1.462 | 0.523 | 0.195-1.404 | 0.456 | 0.158-1.314 |
| DPP-4i only user | 133 | 130 | 3 | 0.454 | 0.132-1.566 | 0.460 | 0.131-1.612 | 0.491 | 0.136-1.771 | 0.232 | 0.057-0.954 |
| DPP-4i and RAS user | 130 | 124 | 6 | 0.952 | 0.369-2.452 | 0.786 | 0.299-2.068 | 0.515 | 0.174-1.523 | 0.251 | 0.074-0.850 |

\* Model 1: non-adjusted; \*\* Model 2: adjusted for age and sex; \*\*\* Model 3: adjusted for factors in Model 2 and comorbidity; \*\*\*\* Model 4: adjusted for factors in Model 3 and medications.
DPP-4 indicates dipeptidyl peptidase-4; RAS, renin-angiotensin system; ICU, intensive care unit; OR, odds ratio; CI, confidence interval.

The fractions of those who received intensive care or died were 3.98% and 4.16% among RAS-blockade users and non-users, respectively (Table 3). The OR of severe/lethal cases among RAS-blockade users was 0.954 (95% CI, 0.471–1.933). The aOR, considering the effects of various clinical variables, was also not significant, at 0.599 (95% CI, 0.251–1.431).

### Combination effect of DPP-4i and RAS blockade

To evaluate the synergistic effect of the combination of these two drugs, subjects were divided into 4 groups according to whether DPP-4i and RAS blockade were used, and further analyzed. Their clinical characteristics are summarized in Table S1 in the Supplementary Appendix.

Compared to that of subjects who used neither DPP-4i nor RAS blockade, the aOR was 0.456 (95% CI, 0.158–1.314) for severe/lethal cases of RAS-blockade-only users, 0.232 (95% CI, 0.057–0.954) for DPP-4i-only users, and 0.251 (95% CI, 0.074–0.850) for subjects using both DPP-4i and RAS blockade.

## Discussion

In this study, we compared the characteristics and clinical outcomes of COVID-19 infection upon administration of DPP-4i and RAS blockade, using a large database representative of the Korean population. The aOR for severe/lethal cases of DPP-4i users was 0.362 (95% CI, 0.135–0.971), and that for RAS-blockade users was 0.599 (95% CI, 0.251–1.431). The aOR for the subgroup of those who used both drugs was 0.251 (95% CI, 0.074–0.850), which was not significantly different from that of DPP-4i-only users. These results suggest that the use of DPP-4i for people with DM Is associated with a better clinical outcome of COVID-19 Infection, suggesting that the synergistic effect of the combination of these two classes of drug is not likely to be noticeable. To our knowledge, these results are new facts that have not been reported previously.

Our findings are consistent with the existing experimental research hypothesis that DPP-4i may reduce the severity of COVID-19 because DPP-4 acts as a receptor for coronavirus^6^’^18^. This hypothesis has not been confirmed in large population groups, and our findings may be among the important evidence that supports it. In our study, DPP-4i users were more likely to exhibit comorbidities than non-users, and thus were more likely to be taking medications of different classes. Despite these differences in characteristics, it is interesting to note that the use of DPP-4i is associated with better clinical outcomes of COVID-19 patients with DM. In particular, since DPP-4i rarely causes hypoglycemia when used alone, it is necessary to examine the possibility of this drug being used as an immunomodulator for systemic infections, and not solely as a diabetes drug^19,20^.

It should also be noted that the number of users of SGLT2i was significantly higher among DPP-4i non-users. A large-scale clinical trial of SGLT2i has demonstrated that this drug has a significant effect on the prevention of cardiovascular disease^21,22^. Based on the results of this study, a randomized trial was recently performed to confirm the effect of SGLT2i on COVID-19 patients^23^. However, SGLT2i is likely to have a negative effect on patients with acute phase infections, for which it is important to maintain stable hemodynamics^24,25^. Currently, we are conducting a study on the clinical impact of SGLT2i on COVID-19 patients; using the results, we plan to provide partial answers to important clinical questions as soon as possible.

Since ACE2 is also a known as a physiologic regulator of human coronavirus infection, the possibility that RAS blockade may negatively affect the susceptibility and severity of COVID-19 infection has been hypothesized^26,27^. However, it is known that RAS blockade has anti-inflammatory and antioxidant pleiotropic effects, and research results on preventing pulmonary complication in vulnerable patients have been reported^28,29^. A systemic review states that there is currently no clear evidence as to whether angiotensin converting enzyme inhibitor or angiotensin receptor blocker needs to be started or stopped for viral infections, including COVID-19^15^.

In our study, RAS blockade tended to decrease the aOR for severe/lethal cases; however, this decrease was statistically insignificant. This implies that RAS-blockade users are also more likely to exhibit comorbidities than non-users, and hence, it is possible that this is a vulnerable clinical condition that requires more medication. It is also possible that the number of subjects studied was not sufficient to confirm statistical significance. This may have been because the effects on the long-term clinical course and prognosis were not considered; owing to the limitation of our study design, only the short-term clinical outcomes of the subjects were analyzed. The synergistic effect of the combination of DPP-4i and RAS blockade could not be explained for the same reason.

This study has other limitations. Firstly, since this study was based on claim data, it was difficult to obtain detailed clinical information about the patients. In particular, it was difficult to evaluate the patient’s demographic characteristics, laboratory tests, images, and detailed clinical course information. Secondly, the study results demonstrated that DPP-4i users exhibited better clinical outcomes; however, these results did not imply a clear causal relationship. Thirdly, it was difficult to predict the long-term clinical course and prognosis of the patients because our analysis was only for short-term clinical outcomes. Fourthly, this study may be difficult to generalize with respect to other countries as it was conducted in Korea, where the proportion of severely ill patients and fatalities related to COVID-19 is lower than that in other countries. Lastly, further research is needed because a clear mechanism for explaining the facts obtained in this study has not been established. These limitations necessitate careful interpretation and generalization of the results.

However, this study was based on a large dataset representative of the entire population of Korea, and such large-scale studies are rare. Moreover, with regard to situations where effective prevention or treatment of COVID-19 infection has not been established, our research results contain meaningful data on drugs that are commercially available and applicable to many people. In particular, using Korea’s national medical insurance claim data for the past 5 years, it was possible to consider the Impact of comorbidities and various medications in the analysis.

Currently, efforts are being made in Korea to increase the size and quality of the COVID-19 database used in research for public interest. In the future, detailed observations of the clinical courses of subjects will enable a more detailed understanding of the disease, and It is expected that a more effective methodology for prevention and treatment will be established soon.

In conclusion, this population-based study suggests that DPP-4i is significantly associated with a better clinical outcome of COVID-19 infection. However, the effect of RAS blockade is not significant.

## Data Availability

Currently, in Korea, medication usage data for the past 5 years for those who have claimed a confirmatory test of COVID-19 are managed as a separate database for research. To protect personal information, the export of raw data is strictly prohibited by law, and research is being performed in a way that provides de-identified results when researchers submit program codes for analysis.

https://hira-covid19.net/

## Acknowledgements

Since the COVID-19 database of the National Health Review and Assessment Service of Korea is provided for public interest, it is a principle that all research results and publications be released to open access.

The authors would like to thank Professors Jeong-Taek Woo and Young Seol Kim of Kyung Hee University for their exceptional teaching and inspiration, which encouraged us to conduct the study.

## Author contributions

SYR established the hypotheses, designed the study, and wrote the manuscript. JL, HN, and D-SK were in charge of data analysis. DJK reviewed the results and participated in the discussion. SYR is the guarantor of this work and, as such, has full access to all the data in the study and takes responsibility for the integrity of the data and the accuracy of the data analysis.

